# Development of a novel risk prediction tool for emergency department patients with symptoms of coronary artery disease: A research study protocol

**DOI:** 10.1101/2024.12.20.24319105

**Authors:** Andrew D. McRae, Aysha J. Macci, Jessalyn K. Holodinsky, Tolulope Sajobi, James E. Andruchow, Bjug Borgundvaag, Steven Brooks, Ivy Cheng, Saswata Deb, Patrick Fok, Peter Kavsak, Michelle M. Graham, Jacques Lee, Shelley L McLeod, Frank Scheuermeyer, Venkatesh Thiruganasambandamoorthy, Hana Wiemer, Justin W. Yan, Corinne M. Hohl, the Canadian Emergency Department Research Network (CEDRN) and the Network of Canadian Emergency Researchers (NCER)

## Abstract

Patients with chest pain and symptoms of acute coronary syndromes (ACS) account for over 600,000 emergency department (ED) visits annually in Canada. Over 80% of these patients do not have ACS, and most are discharged from the ED after a thorough evaluation. However, a large proportion of these patients are referred for outpatient objective cardiac testing after ED discharge, even though their short-term risk for major adverse cardiac events (MACE) such as death, new myocardial infarction or need for revascularization is very small. This contributes to substantial low-value healthcare utilization, and limits access for those patients who are more likely to benefit from objective testing.

Existing risk prediction tools were developed prior to the advent of high-sensitivity cardiac troponin assays, were derived in non-representative populations and, when applied to ED patients with low cardiac troponin concentrations, systematically overestimate short-term risk of (MACE).

This multicenter prospective cohort study will enrol ED patients with chest pain to derive and validate a novel risk prediction tool to accurately identify patients at low risk of MACE and not requiring additional cardiac testing from patients who are likely to benefit from additional cardiac testing. We will enroll 6500 patients at 13 Canadian EDs nd prospectively follow them for 30 days to ascertain a primary outcome of MACE. The risk prediction tool developed in this project will guide safe, efficient, appropriate referrals of ED patients with chest pain.

## Background

Chest pain and symptoms of coronary artery disease are a common cause for presentation to an emergency department (ED), accounting for over 600,000 ED visits annually in Canada^1,2^. The key clinical question addressed in the ED is whether these patients have acute myocardial infarction (MI), which is associated with a 30-day mortality risk of 6.1%^3^. Therefore, patients presenting to the ED with chest pain and symptoms of possible MI undergo extensive investigations, including clinical evaluation and testing with electrocardiograms and blood cardiac troponin measurement. Among patients evaluated in the ED for chest pain, up to 10% are diagnosed with MI and admitted to the hospital. Another 5% are admitted for investigations because of concern for unstable angina, a syndrome of chest pain or other symptoms associated with critical coronary stenosis that does not have evidence of acute biochemical injury (i.e, cardiac troponin concentration below the upper reference limit)^2,4,5^. The remaining 85% of patients, who have had MI and other high-risk diagnoses ruled out in the ED, have a short-term risk of major adverse cardiac events (MACE: Death, MI or revascularization) of 2.5% or less^4,5^. Recent guidelines reinforce that patients at low risk of MACE need not undergo additional testing to diagnose coronary disease at the time of their initial hospital encounter^5^. However, in Canada, as many as 40% of these patients are referred for outpatient cardiac testing after ED discharge to screen for undiagnosed coronary disease^5–7^. In the United States, up to 70% of patients at low risk of MACE undergo additional testing, often as inpatients^5,8–11^. This is an ineffective use of healthcare resources. This low-value testing pattern is driven by reliance on outdated risk scores that overestimate risk of MACE, or use of “gut feeling” estimation of pre-test probability of coronary disease, based on the presence of traditional coronary disease risk factors.

Existing risk scores were developed prior to the widespread availability of high-sensitivity cardiac troponin assays^12^, were derived in stable outpatients^13,14^ or undifferentiated patients with chest pain who had not yet had MI ruled out in the ED^15^, include outcomes of MI on the index ED encounter^15^, and do not account for risks associated with sex and/or gender, nor the risk associated with previously-diagnosed coronary disease. For this reason, they overestimate risk among patients who have had MI ruled out using high-sensitivity cardiac troponin assays^12,14^, contributing to over-use of non-invasive cardiac testing after an initial ED evaluation for chest pain.

Our objective is to derive and internally validate a novel risk prediction tool to accurately predict 30-day MACE among ED patients with chest pain in whom MI has been ruled out and are being considered for ED discharge. This risk prediction tool will guide rational, cost-effective decisions around which patients should be referred for additional cardiovascular testing after a thorough ED evaluation for chest pain.

## Methods

We will follow broadly accepted methods for risk score development^16^ to derive and internally validate a novel risk score for prediction of 30-day MACE among ED patients with chest pain who have had MI ruled out, using a prospective, multicenter observational cohort study.

### Setting

Recruitment will take place in the EDs of large academic hospitals across Canada: Foothills Medical Centre, South Health Campus and Rockyview General Hospital, Calgary; London Health Sciences Centre (University and Victoria Campuses), London; Kingston Health Sciences Centre, Kingston; Sinai Health, Toronto; Sunnybrook Hospital, Toronto; the Ottawa Hospital (Civic and General Campuses), Ottawa; Vancouver General and St. Paul’s Hospitals, Vancouver; QEII Health Sciences Centre, Halifax. Each of these EDs has an annual census of over 60,000 patients, including at least 3,000 patients who undergo evaluation for chest pain annually. Consecutive patients will be recruited while research staff are present in the ED, from 8am-8pm, 7 days/week at most sites. ED physicians will be asked to collect data independently of research staff during hours that research staff are not present. Eligible patients will receive usual care provided by the ED clinicians. ECG and laboratory testing will be performed as per usual clinical practice.

This study will leverage the data collection and management infrastructure of the Canadian Emergency Department Research Network (CEDRN). This is Canada’s largest ED research network, initially formed to systematically collect data on patients tested for SARS-CoV-2 at 50 Canadian EDs. CEDRN has accrued data on over 203,000 patients tested for SARS-CoV-2 and its data has been used to develop risk scores for a variety of COVID-related outcomes^17–19^. CEDRN has an established REDCap data management platform and a secure analytic environment.

### Financial Support, Study registration and Ethics Review

This project is supported by a Project Grant from the Canadian Institutes of Health Research (CIHR, PJT-191778).

This study is registered at clinicaltrials.gov (NCT06743672).

The protocol was approved by the University of Calgary Conjoint Health Research Ethics Board (REB-24-0536) with a waiver of the requirement for informed consent. The study will be approved with a waiver of the requirement for consent at all participating institutions.

### Patient population

We will enroll adult patients (age >25) presenting to the ED with chest pain or symptoms consistent with coronary disease, but who do not have an MI or clear alternative diagnosis. Our age cutoff is consistent with prior literature, demonstrating a very low risk of symptomatic coronary disease below age 25.

**Table 1.** Inclusion and Exclusion Criteria.

| Inclusion Criteria | Exclusion Criteria |
| --- | --- |
| 1. Symptoms of chest pain or of suspected cardiac ischemia (eg dyspnea, dyspepsia); | 1. Acute ischemic ECG changes (ST segment elevation or depression, new T-wave inversion, New left bundle branch block; Wellen's/DeWinter T waves); |
| 2. Age 25 or older; | 2. Alternative diagnosis identified in the ED (e.g. pneumonia, pulmonary embolism, aortic dissection, pancreatitis; peri/myocarditis); |
| 3. High sensitivity cardiac troponin concentration below diagnostic criteria for myocardial infarction, as specified in the Fourth Universal Definition of MI<sup>49</sup>; | 3. Acute coronary syndrome or revascularization in previous 30 days; |
|  | 4. Expected lifespan < 6 months per treating clinician. |

### Participant recruitment

Eligible patients will be enrolled using a waiver of consent, consistent with Canadian research ethics guidelines for research that poses only minimal risk to participants^*20*^, to limit selection bias.

### Data collection, variables, and outcomes

Variables to be collected are described in Appendix 1. Symptoms and clinical variables, including pre-test probability of symptomatic coronary disease will be recorded by ED physicians. Data collection from physicians will employ several approaches, depending on site-specific workflows. Sites will use a combination of: 1) Direct data entry into a REDCap data collection tool linked form the site’s electronic medical record or from QR codes posted in the ED; 2) Standardized clinical documentation templates that include all physician-recorded variables; 3) Paper data collection forms to be transcribed into REDCap by site research staff; 4) Staff– or patient-completed data collection forms verified by physicians, with physician completion of specific fields. Demographics, ECG, and laboratory results will be recorded on an ED data collection form completed at the index ED visit by local research staff. Demographics of any missed eligible patients will be collected to verify the absence of selection bias.

### Outcomes

The *primary outcome* for the prediction tool will be the incidence of *MACE* within 30 days after the index ED visit. MACE is defined as all-cause mortality, MI, or revascularization (non-elective coronary bypass grafting or percutaneous coronary intervention). *Secondary* outcomes include individual MACE components: all-cause mortality, MI, and revascularization within 90 days of the index ED visit.

Outcomes will be ascertained first by querying health records and administrative databases of participating hospitals. This approach has been robust in prior studies conducted by this team^21–23^ and will identify most outcomes as the participating sites are the coronary revascularization centers for their geographic region. Outcomes occurring outside of the participating sites will be ascertained by linking to provincial vital statistics, Discharge Abstract Database (DAD) and National Ambulatory Care Reporting System (NACRS) databases via the Canadian Institute of Health Information) Cardiovascular outcome identification algorithms using these administrative data sources have been previously validated and used in prior studies^7,24,25^. All outcomes will be centrally adjudicated by a committee comprised of cardiologists and emergency physicians disagreements settled by majority.

### Data flow and management

CEDRN’s REDCap-based data management platform, housed on a secure research environment at PopDataBC, will be used for data management and analysis. All data will be either directly entered by physicians or entered by research staff at participating sites into an electronic case report form developed using the REDCap electronic data capture platform.

After data quality assurance has been completed within the REDCap database, the data will be transferred to a secure research environment housed at PopDataBC. This secure research environment will be accessible only to study analysts with an electronic access key and will be used as the analytic platform.

No participant identifiers will be transferred to the REDCap database. Each site will retain a master study list containing the participant’s unique study ID and provincial health number for linkage.

Sites will transfer this master list to the CEDRN Provincial coordinating site (Vancouver General Hospital/UBC, Foothills Medical Centre/University of Calgary, Kingston General Hospital/KGH). This will then be sent securely to CIHI for linkage to administrative datasets. CIHI will then securely transfer de-identified outcome data (containing only the participant’s unique study ID) to the PopDataBC secure research environment.

**Figure 1:**
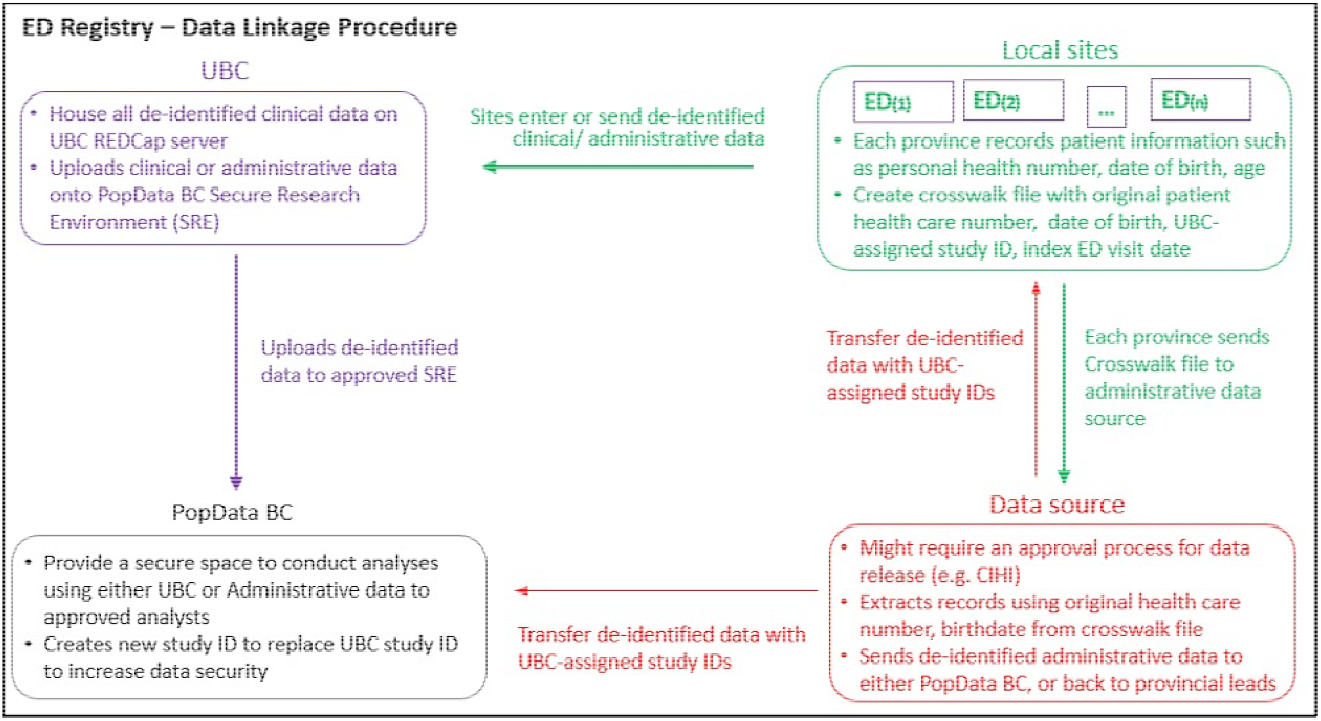
CEDRN Data flow and linkage to administrative data

### Analysis: Prediction rule derivation and internal validation

Descriptive statistics (means, standard deviation, median, percentiles, frequency, and/or percentages) will be used to summarize patients’ demographic and clinical characteristics. Patients’ demographic and clinical characteristics will be used to predict patients’ probability of having 30-day MACE.

For continuous predictors, restricted cubic splines and loess functions will be used to assess the most appropriate functional form of the relationship between each continuous variable and the MACE outcome. Hs-cTn will be modelled in different ways (e.g., absolute value, multiples of limit of quantification, proportion of 99^th^ percentile (i.e., upper reference limit) rather than absolute concentration, absolute/relative change from baseline). Team members have previously developed analytic approaches that can combine different hs-cTn assays to develop a prognostic tool^26^. The association of each functional form of troponin variable with the outcome quantified, and the troponin variable having the strongest association with the outcome will be included in the multivariable model. To prevent overfitting due to large numbers of candidate variables, and to develop a risk score that can easily be used at the bedside, some variables may be combined into composite predictors. For example, rather than including all coronary risk factors in a model, we will create a composite categorical variable indicating the presence of three or more coronary risk factors as opposed to including each individual risk factor in the model. Rather than including all granular symptom variables, we may create a composite “classical anginal symptoms” variable. These will be created with expert clinician input to ensure sensibility and utility to end-users.

The MACE risk prediction model will be developed using a logistic regression model that includes patients’ demographic and clinical characteristics as candidate predictors. Current evidence suggests that regression models have comparable predictive accuracy to machine learning algorithms for risk prediction^27,28^ and support the development of an interpretable integer based score. To fit the most parsimonious set of predictors which maximizes the model’s predictive accuracy the LASSO method will be used. LASSO is a penalized estimation technique which simultaneously achieves variable selection and coefficient shrinkage to mitigate overfitting^29^. Through this selection and shrinkage process, LASSO can also effectively handle collinearity. Interactions between sex and other predictor variables will be quantified and, if indicated, sex-specific models will be created.

The discriminatory performance of the model will be evaluated using sensitivity, specificity, the area under the receiver operating characteristic curve (AUC), and F1 score. To account for the imbalance in the dataset, the classification of each individual based on logistic regression will be weighted using the inverse of the ratio of the size of the “Event” to the “No Event” group as the penalty for misclassification of each individual. For example, the cost of misclassifying a patient at high risk of MACE in 30 days will be higher than the cost of misclassifying an individual with small risk of MACE. The calibration performance of the model will be derived using Brier score and graphically. In the latter, we will graphically compare predicted and observed proportions of the outcome across deciles of model predicted probabilities.

To internally validate the 30-day MACE risk model, repeated 10-fold cross validation will be used. This will be done by randomly splitting the data into 9:1 ratio, retraining the model in the nine-tenth of the data, and estimating the accuracy in the one-tenth. This process will be repeated 10 times. The final accuracy of the model will be obtained as the mean discrimination and calibration measures (AUC, sensitivity, specificity, F1 score, Brier score). We will externally validate the trained model in a temporally and geographically distinct cohort of eligible patients in a future study.

An integer-based risk score will be created from the prediction model by generating regression coefficient-based point scores^30,31^. This will simplify the estimation of a patient’s risk by assigning integer values to each level of each predictor, allowing clinicians to easily estimate risk by summing integers. The discrimination and calibration of the integer-based score will be evaluated as above, and sensitivity and specificity at different score cut-offs will be quantified.

We will compare the performance of the novel risk score (sensitivity, specificity, proportion of patients classified as low risk) to existing risk scores using net reclassification indices and decision curve analyses. We will compare observed physician referral patterns to expected referral patterns based on the integer-based risk score. We will also conduct sensitivity analyses stratifying sites by outpatient testing rates to identify any differences in outcome rates attributable to testing patterns and will conduct secondary analyses to validate the risk score in samples from high-vs. low-referral rate sites. All analyses will be conducted using Python software (Version 2.7, available at www.python.org) and R (R-project.org). PopDataBC integrates RStudio for analysis within its secure research environment.

### Sample Size and precision

Current risk scores, when used in conjunction with hs-cTn testing, have a specificity no better than 60%^12^. Our goal is to develop a score with a specificity of at least 70%. This analysis is designed to maximize specificity while maintaining a fixed sensitivity. We propose a target sensitivity of 98.5% for a dichotomized score, as this is the standard to which other risk scores have been developed^32,33^ and approximates emergency physicians’ documented risk tolerance of 1-2% for missed cardiac events^34^. Data from a 1184 patient study from our lead site^35,36^ suggests that the 30-day MACE risk among patients with MI ruled out using high-sensitivity troponin is approximately 2.5%. We will conservatively estimate the 30-day MACE risk in our cohort at 2% for the purposes of sample size calculation, which will require a large cohort to ensure acceptable precision around our target sensitivity. A sample of 6350 patients, with 127 events, would give an acceptable precision around a target sensitivity of 98.5% for a dichotomized risk score. The study will be overpowered for specificity, with excellent precision around a range of specificity point estimates. Appendix 2 details precision around various combinations of sensitivity and specificity with different permutations of sample size and 30-day MACE risk.

## Discussion

### Feasibility, challenges, and solutions

Participating sites are using different high-sensitivity troponin assays (Appendix 3), which will enhance the generalizability of our work. This will not lead to variation in eligibility by site, as patients are eligible if they do not have an MI as defined by the 4th Universal Definition of MI, with specific diagnostic cutoff concentrations for each available hs-cTn assay^37^. We have previously developed analytic approaches that can combine different hs-cTn assays for prognostication^26^. Because we are excluding patients with MI on the index ED visit, losses in specificity owing to using a more sensitive biomarker will be minimized.

The primary outcome, MACE, will be driven largely by the need for revascularization. Incident MI and mortality are rare in this population. Revascularization, as an outcome, has been criticized as subjective^38^. However, revascularization is an important patient-oriented, clinically relevant and system-level outcome that generally only occurs in patients with symptomatic, angiographically proven high-grade coronary disease. In Canada, there are no financial incentives for institutions to perform revascularization procedures without clinical indications.

We expect referral patterns, timing and modalities of post-discharge investigations to vary between sites, which will enhance the generalizability of our work. We will use sensitivity analyses to evaluate whether referral practices influence outcome incidence. However, based on our experience, patients requiring urgent revascularization will be identified within 30 days of an index ED visit at all sites based on standard care.

It is unlikely that the derived risk score will have suboptimal discrimination as measured by the c-statistic. Even if this were to occur, the c-statistic does not necessarily correlate with clinical utility, which is the most important factor that guides decision-making. A score needs to perform well with high sensitivity and specificity at specific cutoff values. We will quantify the sensitivity and specificity at different score cut points, compare the predicted referral pattern for each patient directed by the score to observed practice and existing risk scores, and estimate potential impact of score use on clinical care.

### Future Research Directions

We envision several secondary uses for these data. We will use these data to quantify variation in referral and testing practices between sites to facilitate targeted knowledge translation interventions to reduce low-value testing. These data will also be used to develop a base case for economic evaluations to be conducted in future validation and implementation studies. Although regression-based approaches are superior to machine learning for generating risk prediction tools^27,28^, as machine learning algorithms to not generally lead to user-friendly models that can be easily applied at the bedside, we will nonetheless use these data for exploratory analyses to develop machine learning algorithms to predict cardiovascular outcomes.

After completing the derivation and internal validation of this risk score, we will seek separate grant support for a prospective, multicenter external validation study. We will seek separate grant support to biobank patient blood samples to update this risk score as novel biomarkers become available.

## Conclusions

Chest pain and symptoms of coronary disease are among the most common reasons for ED visits in Canada. Rational use of outpatient cardiac testing among ED patients with chest pain who have had MI ruled out remains a challenge. Using the infrastructure of Canada’s largest emergency medicine research network, and extensive experience developing risk prediction tools, we will develop a modern risk prediction tool to accurately estimate Canadian patients’ risk of MACE within 30 days of having MI ruled out in the ED. An accurate risk prediction model will guide rational referral patterns that could safely save over 50,000 unnecessary consultations and non-invasive cardiac tests per year in Canada, facilitating optimal healthcare resource utilization, and optimizing patient access to cardiovascular testing resources.

## Data Availability

All data produced in the present study are available upon reasonable request to the authors

