## Appendices for "Development of a novel risk prediction tool for emergency department patients with symptoms of coronary artery disease: A research study protocol"

Appendix 1. List of variables

| Variable | Values | Variable type | Collecting/recording individual |
|---|---|---|---|
| **Identification** | | | |
| Site | FMC, SHC, VGH, StP, LHSC, SREMI, OHRI, Queen's | | Research staff or recorded by direct REDCap entry |
| Date of Presentation | Date | | Research staff or recorded by direct REDCap entry |
| Time of presentation | Time | | Research staff or MD |
| Time of initial physician assessment | Time | | Research staff or MD |
| **Demographics** | | | |
| Patient initials | Freetext | Discrete | |
| Age in years | 18- | Continuous | Research staff or MD |
| Biological Sex | Male, Female | Binary | MD |
| Gender (if different from biological sex) | man, woman, oonbinary, other | Categorical | MD |
| Forward Sortation Area | First 3 digits of postal code | Freetext | Research Staff |
| **Clinical Characteristics** | | | |
| Date/time of onset of maximal symptoms | | | MD |
| Pattern of Chest pain | Single Episode/constant<br>Intermittent | Categorical | MD |
| Duration of Longest episode of chest pain | Hours/Minutes | Continuous | MD |
| Character of Chest pain | Pressure<br>squeezing,<br>burning,<br>sharp/stabbing<br>Dull<br>Aching,<br>Other,<br>Tearing | Categorical | MD |
| Provoking factors | exertion,<br>movement, | Categorical | MD |

| | | | |
|---|---|---|---|
| | deep inspiration,<br>positional changes<br>Palpation<br>Eating/drinking<br>stress<br>None<br>Other | | |
| Relieving Factors | positional changes<br>Rest<br>Analgesics<br>Nitroglycerin<br>Eating/drinking<br>Deep inspiration<br>other<br>none | Categorical | MD |
| Radiation | L shoulder/arm,<br>R shoulder/arm,<br>both shoulders/arms,<br>neck/jaw,<br>back/interscapular region,<br>upper abdomen<br>non-radiating | Categorical | MD |
| Reproducibility | Reproduced with palpation | Binary | MD |
| Associated symptoms/signs | dyspnea,<br>N/V,<br>diaphoresis,<br>fatigue,<br>lightheadedness/presyncope<br>palpitations | Categorical | MD |
| CAD risk factors | Hypertension<br>Dyslipidemia<br>Smoking (current or previous)<br>Diabetes Mellitus<br>Family history of premature CAD (Age <65)<br>Prior pre-eclampsia<br>Prior COVID infection | Categorical | MD |
| Prior history of CAD (imaging-documented ischemia/stenosis | Yes/No | Binary | MD |

| | | | |
|---|---|---|---|
| >50%, prior revascularization, patient-reported history of CAD or medical therapy for MI/angina) | | | |
| Classification of chest pain | Highly suspicious for angina/CAD<br>Moderately suspicious for angina/CAD<br>Slightly suspicious for angina/CAD | Categorical | MD |
| ECG variables | Acute coronary occlusion (STEMI or STEMI equivalents eg. Wellen's, Dewinter, new LBBB)<br>Other probable ischemic changes (Dynamic/new STD, New Q waves, or T inversions, perfusing VT)<br>Nonspecific ECG changes<br>Chronic conduction system changes (RBBB/old LBBB/LAFB/LPFB)<br>Symptomatic supraventricular dysrhythmia (Afib, atrial tachycardia, SVT, Junctional tach)<br>Sinus tachycardia<br>Bradydysrhythmia/3rd degree heart block/new AV block<br>Normal | Categorical | MD |
| Troponin testing | | | |
| Date and Time of blood draw for first Troponin | Hours, minutes | | Research staff |
| Troponin 1 concentration | | continuous | Research staff |
| Date and Time of blood draw for first | Hours, minutes | | Research staff |

| | | | |
|---|---|---|---|
| Troponin second Tn test | | | |
| Tn concentration 2 | | continuous | Research staff |
| Date and Time of blood draw for NthTroponin Tn test | Hours, minutes | | Research staff |
| Tn concentration N | | continuous | Research staff |
| **Clinical Impression** | | | |
| Pretest probability of symptomatic coronary disease (new or old) as cause for their presentation? | 0-100% | Continuous/VAS | MD |
| ED diagnosis | Free text | To be grouped into categorical variables | MD |
| ED disposition | Cardiology consult in ED for admission<br>Cardiology consult in ED for opinion<br>Discharge | categorical | MD or research staff |
| investigations/referral ordered by ED physician | Outpatient Cardiologist consultation/internist consultation<br>Exercise ECG test<br>Holter monitor<br>Echocardiogram<br>Stress echo<br>Nuclear scan/MPI<br>Coronary CT<br>Cardiac MR<br>None | categorical | MD |
| **Laboratory Values (from EMR)** | | | |
| RDW | | Continuous | Research staff |

| | | | |
|---|---|---|---|
| Glucose | | Continuous | Research staff |
| eGFR | | Continuous | Research staff |
| NT-proBNP (if done) | | Continuous | Research staff |
| Outcomes | | | |
| 90-day Mortality | Y/N | Binary | Research staff |
| Time from presentation to death | Days | Continuous | Research staff |
| 90-day MI after ED discharge | Y/N | Binary | Research staff |
| Type of MI after ED discharge | Type1/2/3/4<br>SCAD | Categorical | Research staff |
| time from index presentation to Dx of MI | Days | Continuous | Research staff |
| 90-day revascularization (unplanned PCI, unplanned CABG) | Y/N | Binary | Research staff |
| Time from index presentation to revascularization | Days | Continuous | Research staff |

Appendix 2. Precision around estimates of sensitivity and specificity for a dichotomized risk score based on permutations of sample size and 30d MACE rate

| Sample size | 30d MACE Rate | Sensitivity | 95% CI for sensitivity | Specificity | 95% CI for Specificity |
|---|---|---|---|---|---|
| 6350 | 2% | 98.5% | 94.3-99.6% | 30% | 28.9-31.2% |
| 6350 | 2% | 98.5% | 94.3-99.6% | 40% | 38.8-41.2% |
| 6350 | 2% | 98.5% | 94.3-99.6% | 50% | 48.8-51.3% |
| 6350 | 2% | 98.5% | 94.3-99.6% | 60% | 58.9-61.2% |
| 6350 | 2% | 98.5% | 94.3-99.6% | 70% | 68.9-71.2% |
| 5100 | 2% | 98.5% | 93.4-99.5% | 30% | 28.8-31.2% |
| 5100 | 2% | 98.5% | 93.4-99.5% | 40% | 38.7-41.2% |
| 5100 | 2% | 98.5% | 93.4-99.5% | 50% | 48.7-51.3% |
| 5100 | 2% | 98.5% | 93.4-99.5% | 60% | 58.9-61.2% |
| 5100 | 2% | 98.5% | 93.4-99.5% | 70% | 68.9-71.2% |
| 7450 | 2% | 98.5% | 94.7-99.5% | 30% | 28.9-31.1% |
| 7450 | 2% | 98.5% | 94.7-99.5% | 40% | 38.8-41.2% |
| 7450 | 2% | 98.5% | 94.7-99.5% | 50% | 48.8-51.2% |
| 7450 | 2% | 98.5% | 94.7-99.5% | 60% | 58.9-61.2% |
| 7450 | 2% | 98.5% | 94.7-99.5% | 70% | 68.9-71.2% |
| 6434 | 3% | 98.5% | 95.4-99.5% | 30% | 28.9-31.2% |
| 6434 | 3% | 98.5% | 95.4-99.5% | 40% | 38.8-41.2% |
| 6434 | 3% | 98.5% | 95.4-99.5% | 50% | 48.8-51.3% |
| 6434 | 3% | 98.5% | 95.4-99.5% | 60% | 58.9-61.2% |
| 6434 | 3% | 98.5% | 95.4-99.5% | 70% | 68.9-71.2% |

**Appendix 3. hs-cTn assays in use by participating site**

| Site | Assay | 99th percentile | Limit of quantification | 10% Coefficient of variation |
|---|---|---|---|---|
| Vancouver General Hospital | Siemens Vista hs-cTnI | Female: 51ng/L<br>Male: 76 ng/L | 1ng/L | 10ng/L |
| The Ottawa Hospital (Civic and General Sites) | Abbott ARCHITECT hs-cTnI | 30 ng/L | 1ng/L | 10ng/L |
| Kingston Health Sciences Centre | Abbott ARCHITECT hs-cTnI | 30 ng/L | 10ng/L* | 10ng/L |
| London Health Sciences Centre (Victoria and University Campuses) | Roche Elecsys TnT-hs | 14 ng/L | 3ng/L | 13ng/L |
| St. Paul's Hospital | Roche Elecsys TnT-hs | 14 ng/L | 5ng/L | 13ng/L |
| Mt. Sinai Hospital | Roche Elecsys TnT-hs | 14 ng/L | 3ng/L | 13ng/L |
| Foothills Medical Centre | Roche Elecsys TnT-hs | 14 ng/L | 3ng/L | 13ng/L |
| Rockyview General Hospital | Roche Elecsys TnT-hs | 14 ng/L | 3ng/L | 13ng/L |

* 10ng/L is the lowest reported concentration in clinical practice, however smaller concentrations can be quantified and reported for research studies.
